# A New Mixed Frequency Regression Model For Environmental Epidemiology

**DOI:** 10.64898/2026.06.03.26354801

**Authors:** Nidhi Shukla, Suzanne E. Bartington, Anna L. Hansell, Tim CD Lucas

## Abstract

**Background:** In the absence of high-resolution response data, exposure-response modelling often relies on aggregated low-frequency exposure data, leading to loss of high-resolution information. Mixed Data Sampling (MIDAS) from econometrics offers an alternative but is limited due to its inability to make high-resolution predictions, inflexible likelihoods and penalised nonlinear functions, and limited visualization options. We propose a mixed-frequency Distributed Lag Non-linear Model (mf-DLNM) which can eliminate the need to aggregate exposure data in environmental epidemiology and provide high resolution predictions for time series studies.

**Methods:** We evaluated the inference and predictive performance of the mf-DLNM. To evaluate its ability to estimate exposure–response relationships, we applied mf-DLNM and same-frequency (sf)-DLNM using data from the West Midlands, UK. Additionally, we compared the predictive performance of mf-DLNM with sf-DLNM and MIDAS across nine regions of England. As MIDAS cannot predict at the resolution of the predictor (daily), we compared the predictive performance of mf-DLNM and MIDAS at weekly resolution. To test the model’s ability to predict high temporal resolution risk (daily), we compared sf-DLNM (with access to daily mortality counts) with mf-DLNM (with access only to weekly mortality counts).

**Results:** In the West Midlands example, mf-DLNM performed comparably to sf-DLNM in estimating daily risk of temperature on respiratory mortality. Furthermore, mf-DLNM and MIDAS exhibited similar performance for weekly predictions. For high-resolution predictions, mf-DLNM and sf-DLNM showed nearly similar performance, despite mf-DLNM having access only to low-resolution response data.

**Conclusion:** This mixed-frequency approach in environmental epidemiology overcomes the limitations of predicting health risks using aggregated exposure data and provides estimates of high-resolution outcomes in the absence of high-frequency health outcome datasets.

## 1. Introduction

Extreme environmental events are known to pose health risks for vulnerable human populations.^1^ Extremes of temperature have been widely associated with mortality in various studies.^2–4^ Non-optimal temperatures have been associated with 9% of total global deaths, computed based on the location-specific minimum mortality temperature.^5^ Additionally, in 2019, chronic respiratory diseases ranked as the third leading cause of mortality, accounting for 4 million deaths globally, where non-optimal temperatures were one of the prominent risk factors.^6^ Some studies have estimated the hourly impact of environmental exposure to temperature on human health using outcomes such as ambulance calls for all-cause or cause specific diseases.^7–9^ However, these studies are rare due to the limited availability of routine health outcome data at higher temporal resolution. While outcome data are often available only at a low resolution (e.g., monthly or weekly), exposure data (e.g., daily or hourly) are often available at higher temporal resolution. Therefore, most exposure modelling studies use aggregated exposure data. We aimed to develop a mixed-frequency model to obtain the relationship between low-temporal resolution health outcomes and high-temporal resolution exposure data and predict a high temporal-resolution health risk using routine data in England.

The concept of mixed-frequency modelling has been widely developed in the field of econometrics.^10^ Econometrics is a domain within economics that uses statistical methods to model relationships between economic observations.^11^ The Mixed Data Sampling (MIDAS) regression model was developed in econometrics for the aforementioned situation where researchers need to aggregate high-resolution information to a lower resolution. Thereafter, researchers have widely used MIDAS in econometrics for various situations, including forecasting economic growth and stock returns.^12–14^ While MIDAS has been used to establish relationships between mixed-frequency data, the R package for MIDAS is “midasr”. Additionally, the MIDAS model currently only employs the normal likelihood, while Poisson and negative binomial distributions are crucial in environmental epidemiology. Furthermore, it has limited visualisation functionality for exposure-response modelling. For similar situations where high-frequency exposure data are available for low frequency response data, a recent study applied DLNM employs a disaggregation method to obtain the relationship between high-frequency exposure with the low-frequency response^15^. In contrast, this study adopts a mixed-frequency modelling framework for such scenarios, offering a more flexible and familiar approach.

Considering the limitations of MIDAS and its unfamiliarity among researchers in environmental epidemiology, we propose using the distributed lag nonlinear model (DLNM), which is an extension of the distributed lag model (DLM) ^16,17^ for mixed-frequency exposure modelling. The concept of DLM was first proposed by Koyck ^18^, as a geometric or infinite distributed lag model, while a more flexible and stable method for estimating the lag effect known as the polynomial or the finite distributed lag model, was developed by Almon^19^. In the DLM model the lagged weights are approximated by a polynomial of suitable degree. Early studies used DLM in environmental epidemiology to examine the association between environmental factors and mortality.^20,21^ However, the DLM cannot model non-linear exposure-response relationships which is important in environmental epidemiology.^22,23^ Therefore, DLNM, which is an extension of DLM, has been developed in environmental epidemiology.^16^ The DLNM framework has been extensively used in exposure-response modelling for describing the potential non-linear delayed effects of environmental exposures.^24–27^

To understand the rationale for using DLNM in mixed frequency modelling, it is important to comprehend the similarities and differences between DLNM, its precursor DLM, and MIDAS. MIDAS was developed in econometrics based on the concept of DLM, while DLNM was developed in environmental epidemiology using the same foundation.^10,16,19^ Therefore, in principle, these models are similar. Specifically, both DLM and MIDAS typically use polynomial functions to model predictor-response relationships over time ^28^, therefore the parametric function that handles lag is the same in both. The main distinction between MIDAS and DLM lies in the frequency of predictors and response. MIDAS is a mixed-frequency model, whereas DLM applies on same-frequencies. MIDAS handles mixed frequencies by using a predictor matrix of the lagged values of high-frequency variables. Thus, it is possible to adapt the DLM framework to capture mixed-frequency relationships using a similar approach. However, as previously mentioned, DLM lacks the features necessary to capture potential non-linearities in exposures-response relationships. Consequently, we propose using DLNM to develop mixed-frequency exposure-response relationships, given its ability to model non-linear and delayed effects of exposures and response and its effective visualization capabilities.

We implemented the mf-DLNM and conducted an example analysis using data from the West Midlands, UK, to estimate the relationship between mixed frequency exposure (daily temperature) and response (weekly deaths). The results were compared with the estimates from the same-frequency (sf-) DLNM, which was fitted to daily temperature and daily mortality data. To further evaluate the applicability of mf-DLNM, its predictive performance was assessed relative to both MIDAS and sf-DLNM. Given that MIDAS cannot make predictions at the resolution of the predictor (daily), we tested the performance of mf-DLNM against MIDAS in making predictions at a weekly resolution (i.e., we tested whether mf-DLNM could predict the relationship with mixed frequency data). We then compared mf-DLNM to sf-DLNM, focusing on the predictive performance of daily health outcomes. In this comparison, we established the capability of mf-DLNM to predict high temporal resolution risk (daily) using low-resolution health data. We conducted this detailed methodological assessment using respiratory mortality and temperature data from nine regions of England.

## 2. Method

The mf-DLNM was compared with the sf-DLNM and MIDAS models in terms predictive performance for nine regions of England. Additionally, its ability to characterize the exposure–response relationship was evaluated by comparing the relative risks (RR) estimated by the mf-DLNM and the sf-DLNM for the West Midlands region. The mf-DLNM used aggregated weekly mortality data, whereas the sf-DLNM was fitted using daily mortality data.

The mixed-frequency model developed for environmental epidemiology was primarily designed to enable statistical inference on exposure–response associations. However, the results presented in this study also evaluated the model’s predictive performance to compare it with traditional approaches such as MIDAS. We note that assessing a models predictive performance on real-world data is objective and interpretable. In contrast, comparing the parameter estimates of two fitted models is less interpretable as neither estimate is objectively true. The application of the model in assessing the temperature–mortality relationship using lower-frequency health data with high frequency exposure data was demonstrated and compared through an example for the West Midlands. It is important to emphasize that this risk assessment was conducted solely for methodological evaluation and is not intended to inform or guide policy development. **Supplementary Figure 1** illustrates the workflow used in this study to test mf-DLNM for environmental epidemiology.

### 2.1 Temperature Data

For our exposure variable, we used ERA5-Land climate reanalysis temperature data from the European Centre for Medium-Range Weather Forecasts (ECMWF) Copernicus programme for January 01, 2010 to December 31, 2020. This temperature data is freely available at an hourly temporal resolution and at ~9 km of spatial resolution (https://climate.copernicus.eu/climate-reanalysis). The data have been assessed for usability and reliability and have been used in various temperature-mortality relationship studies.^29–31^ Gridded ERA5 data were aggregated by regions to obtain region-specific daily temperature time series for the study period.

### 2.2 Respiratory mortality Data

Extreme heat and cold temperatures have shown a significant positive association with respiratory mortality, among other causes, in England and Wales.^32,33^ Therefore, for health outcomes, we used daily mortality counts during January 01, 2010 to December 31, 2020, due to diseases of the respiratory system, defined using the International Classification of Diseases, Tenth Revision (ICD-10) codes J00–J99. The data was obtained from the Office for National Statistics (ONS) (https://www.ons.gov.uk/). To replicate the common situation in which only low temporal resolution data are available, we aggregated the mortality counts by week.

### 2.3 Overview of MIDAS and DLNM statistical models

MIDAS is a time series model where the data involved are at different frequencies.^10^ It has been widely used in macroeconomics and finance, where the explanatory variables are present in high frequency and variables of interest are only available in low frequency. Statistically, MIDAS is similar to DLM, except that it handles low-frequency response data and high-frequency predictors rather than using the same frequencies as in DLM. MIDAS handles mixed frequencies through a predictor matrix formed from the lagged values of the high-frequency predictors. A general model representation to describe MIDAS is given by

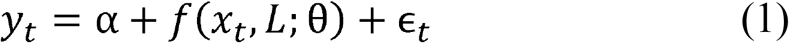

where y_t_ is the response variable at low frequency at time t. x_t_ is the predictor variable at high frequency at time t, which comes from a vector of lagged values of the predictor variable, given as 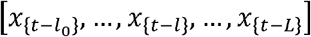. ℓ_0_ and L is the minimum lag and maximum lag, respectively. Here, if y_t_ is at weekly resolution and x_t_ is daily resolution, then L will be 7. Further, *α* is the intercept and *ϵ*_t_ represents normally distributed residuals.

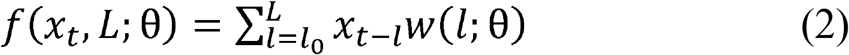

The parameterization of the lagged coefficients typically uses a finite polynomials function given by w(l; *θ*) and *θ* is parameters of lag weighting function (shape of lag weights).^34^ This has come from Almon lag, which has been conceptually used in the estimation of distributed lag models.^17,21^ The two different approaches for constructing lag polynomials are the normalized (“nealmon”) and non-normalized (“almonp”) exponential Almon lag polynomials. Moreover, there also exists an alternative version, such as the non-parametric MIDAS model.

The non-parametric MIDAS employs a penalized least-squares estimator, which introduces a certain degree of smoothness to the distribution of lags.^35^

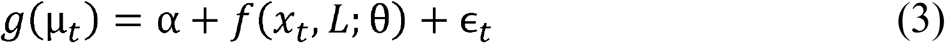

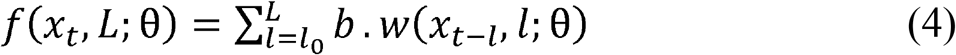

The function b.w(x_t-l_, l;*θ*) combines an exposure-response function b(x) and a lag-response function w(l), expressing a bi-dimensional basis function called the cross-basis which is a product of the basis function of predictor and lag.^36^ Thus, *θ* controls the shape of basis function, which are parameterised through a combination of coefficients denoted as *β*. Although the DLNM model has been extensively developed in environmental epidemiology, it has primarily been applied to same-frequency datasets. However, availability of high-frequency exposure information but low-frequency variables of interest (such as health outcomes) is a common situation in epidemiology. Therefore, this study aims to employ the concept of MIDAS and the framework of DLNM for mixed-frequency regression in environmental epidemiology.

### 2.4 Fitting same and mixed frequency models

We fitted a mixed-frequency model with daily temperature data and weekly mortality count using the MIDAS approach. The MIDAS regression ^10,37^ is given by;

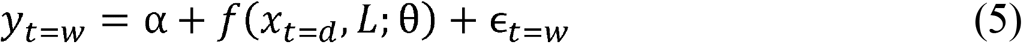

where t is the time of observation, y_t_ represents weekly respiratory mortality count which is the low-frequency dependent variable, x_t_ represents daily temperature which is the high-frequency independent variable, w(l;*θ*) is a chosen function that parametrises the lag structure using a small-dimensional vector of parameters θ and *ϵ*_t_ represents the error term. L represents the maximum lag (i.e., 7 for this case). *α* is the intercept and *ϵ*_t_ represents normally distributed residuals. We used non-parametric MIDAS regression, and smoothing parameters were chosen based on Akaike Information Criterion (AIC).^35^

In addition, we fitted two DLNM models, sf-DLNM and mf-DLNM, using daily temperature as the predictor in both, with daily and weekly mortality as the response variables in the respective models. We built these models using penalized splines within generalized additive models (GAM) with Poisson distribution and log link.^38^ We used the GAM with penalized splines as it provides various advantages, including reduced estimation error and improve predictive accuracy.^39,40^ For comparison, the spline of time was not included in these models because MIDAS does not involve nonlinear spline functions to control for long term trends and seasonality like DLNM. Therefore, controlling for long term trends and seasonality would have provided better predictions for both mf-DLNM and sf-DLNM. Furthermore, in the mf-DLNM, the low-frequency weekly respiratory outcome was aligned to a specific day of the week, such as last day of the week. Therefore, the lag 0 for mf-DLNM will be the final day of the 7 days that the response is aggregated over. In contrast, the sf-DLNM follows the same approach as the standard DLNM, therefore, lag 0 is the day of the response, while lag 1 is the day response data. Furthermore, one key difference in the lag structure between the two models is that, in the sf-DLNM, exposure days are reused across daily outcomes. For example, the lag 1 exposure for day 10 is the lag 2 exposure for day 11. In contrast, in the mf-DLNM, each daily exposure contributes only once to the weekly outcome. For comparison of predictive performance, the three models (MIDAS, mf-DLNM, and sf-DLNM) were fitted with a lag of 7.

For these models, the relationship between temperature and mortality is given as;

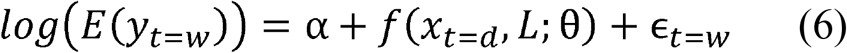

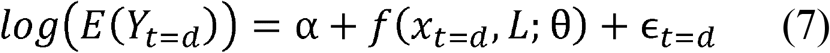

where y_t=d_ represents daily respiratory mortality counts and y_t=w_ represents weekly respiratory mortality counts at time t, x_t_ represents daily temperature at time t, *α* is the intercept and f(x_t_, L; *θ*) is the cross-basis matrix for temperature and lag. As in equation (4) the b.w (x_t-l_, l;*θ*) term is made up of chosen functions that parameterize the non-linear exposure-response relationship with a smaller set of coefficients *θ*. Further, *β* represents the parameters of these functions. The crossbasis was the same for both of these models. We fitted the models for a maximum lag of 7. We chose 6 and 5 degrees of freedom (df) for the basis dimension. The penalization was performed via paraPen in the GAM, which automatically control the smoothness and effective degrees of freedom (edf). The smoothing parameters were estimated using REML. The values of AIC for these models are provided in the **Supplementary Table 1**.

X_p_ is a prediction matrix that contains the basis functions evaluated at the observed predictor values. It contains the observed predictor values transformed into a transformed (basis) space, which simplifies the representation of complex relationships. To obtain the daily prediction 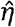 from the mf-DLNM, we multiplied this prediction matrix (X_p_) by the estimated parameters 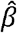. This process, allows us to evaluate each smooth term in the model at the specific predictor values, resulting in daily predicted death counts.^40^ Therefore, by multiplying the basis-transformed predictors (X_p_) with the coefficients 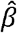, the smooth curves are **reconstructed** at the observed predictor values (daily). These give the predicted weekly values for that day, which are then simply scaled to daily predictions.

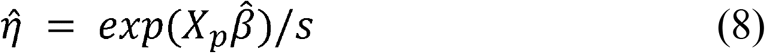

Here, s represents the scaling factor, which is analogous to the maximum lag (i.e. 7 for weekly and daily data).

These models were fitted using R (R Core Team, 2021) with packages dlnm ^17^, mgcv ^41^, and midasr ^37^ and other analysis and visualisation was performed using ggplot.^42^

### 2.5 Demonstrative example analysis using mf-DLNM for West Midlands

We applied mf-DLNM in an example analysis for the West Midlands, UK, to investigate the influence of temperature on overall respiratory mortality from January 01, 2010, to December 31, 2020. The primary objective was to illustrate the application of mf-DLNM with a mixed-frequency dataset and to estimate RR at high temporal resolution using a low-resolution outcome dataset. Additionally, we compared RR estimates between the mf-DLNM and sf-DLNM models. The analysis is based on the model in equation (6) using a GAM. This model used the Poisson distribution and was used to estimate the mixed frequency relationship of exposure and response. It was fitted with a maximum lag of 21 days, chosen following earlier research to capture both immediate and delayed impact of hot and cold weather.^43^ Additionally, a spline of time, with 7 degrees of freedom (df) per year, was used to describe long-time trends and seasonality.

### 2.6 Example using NMMAPS

While the data for the West Midlands example analysis is accessible, we also provide a more user-friendly example analysis using publicly available data that can be easily downloaded. The code for this analysis is available in Supplementary Material (E1). The code reproduces the mf-DLNM for obtaining the relationship between exposure and response using National Morbidity, Mortality, and Air Pollution Study (NMMAPS) data for the period 1987-2000.

### 2.7 Method Performance Analysis

Subsequently, models were fitted for all nine regions of England to illustrate their performance at varying strengths of association. The primary objective was to compare the performance of mf-DLNM to the existing models (sf-DLNM and MIDAS) under varying levels of association. The performance of the models was assessed using the root mean square error (RMSE) and Pearson correlations coefficient (R-values) between observed and predicted mortality counts. Considering, that MIDAS could only predict the dependent variables at low frequency, the comparison between MIDAS and mf-DLNM was performed using weekly observed and predicted deaths. We also computed the RMSE and the correlation coefficient between sf-DLNM and mf-DLNM to evaluate the performance of mf-DLNM in predicting high-frequency response variables along with performing mixed-frequency exposure-response modelling. It is worth noting that the comparison of sf-DLNM and mf-DLNM was not to determine which method is better; given that sf-DLNM was fitted to daily data it is clear that it has more information and will have better performance. Instead, this comparison aims to evaluate how closely the mf-DLNM performs relative to the sf-DLNM, despite having access only to weekly data.

The data and code to implement the study method and analysis in open-source R software is available from https://github.com/nidhienv01/Mixed-Frequency-DLNM.

## 3 Results

### 3.1 Descriptive analysis

Over the period from January 01, 2010 to December 31, 2020, 724,935 cases of respiratory mortality were reported across nine regions in England. More deaths were observed during the winter months compared to the summer months. During this time, the North East and North West had the highest rates, followed by the South West, while London had the lowest rates. **Table 1** provides a region-specific descriptive overview of respiratory mortality and temperature profile.

**Table 1:** Descriptive statistics of respiratory mortality and temperature across regions of England during the study period from January 01, 2010 to December 31, 2020.

| Region | Respiratory Mortality |  | Temperature |  |  |
| --- | --- | --- | --- | --- | --- |
|  | Counts | Crude Rate per 1000 population | Mean | Mean Max | Mean Min |
| West Midlands | 78957 | 13.1 | 9.7 | 23.8 | -7.3 |
| East Midlands | 64383 | 13.2 | 9.9 | 25.1 | -7.3 |
| East of England | 80967 | 12.8 | 10.6 | 25.5 | -4.3 |
| London | 71497 | 8.1 | 10.8 | 26.9 | -4.8 |
| South West | 77699 | 13.6 | 10.5 | 23.5 | -5.7 |
| South East | 117518 | 12.7 | 10.7 | 25.5 | -3.9 |
| North West | 113296 | 15.3 | 8.8 | 22.0 | -8.4 |
| North East | 42818 | 16.2 | 8.3 | 21.5 | -7.5 |
| Yorkshire and The Humber | 77800 | 14.0 | 9.1 | 23.2 | -7.9 |

### 3.2 Example analysis using mf-DLNM Model for West Midlands

We first applied mf-DLNM to the West Midlands region as a demonstrative example of its application and compared the estimated RR between mf-DLNM and sf-DLNM, as outlined in the preceding section. The estimated RR may vary with the inclusion of other potential covariates; therefore, similar demonstrations were not conducted for other regions. Furthermore, it is important to note that this risk assessment was conducted exclusively for methodological evaluation purposes and is not intended to guide policy development. The primary aim was to evaluate the performance of the mf-DLNM approach in the context of mixed-frequency modelling within environmental epidemiology. **Figure 1** depicts the overall cumulative RR of respiratory mortality over a 21-day lag period in relation to daily ambient temperature. A J-shaped relationship between temperature and respiratory mortality was observed, as shown in the figure. This relationship is well established in the literature.^26,44^ Both high and low temperature showed significant risk of increased respiratory mortality in the West Midlands. The overall RR of respiratory mortality, compared to the reference temperature (mean temperature: 10°C), was 1.34 (1.06 –1.71) for extreme cold (0.1st percentile of temperature) and 1.06 (0.84–1.33) for extreme heat (99.9th percentile of temperature). Subsequently, **Figure 2** illustrates the effect of temperature on respiratory mortality at specific temperatures, corresponding to the 0.1st, 5th, 95th, and 99.9th temperature percentiles representing extreme and moderate cold and moderate and extreme heat and at specific lags (0, 3, 15 and 21). Cold temperatures showed no effect at short lag periods of 0 and 3 days, whereas greater effect at longer lag periods of 15 and 21 days. In contrast, hot temperatures exhibited the opposite pattern, with a stronger effect at shorter lags and a declining effect at longer lags. Therefore, the model indicated that extreme heat exerts a more immediate effect compared to cold days, which aligns with existing literature.^43^

**Figure 1.**
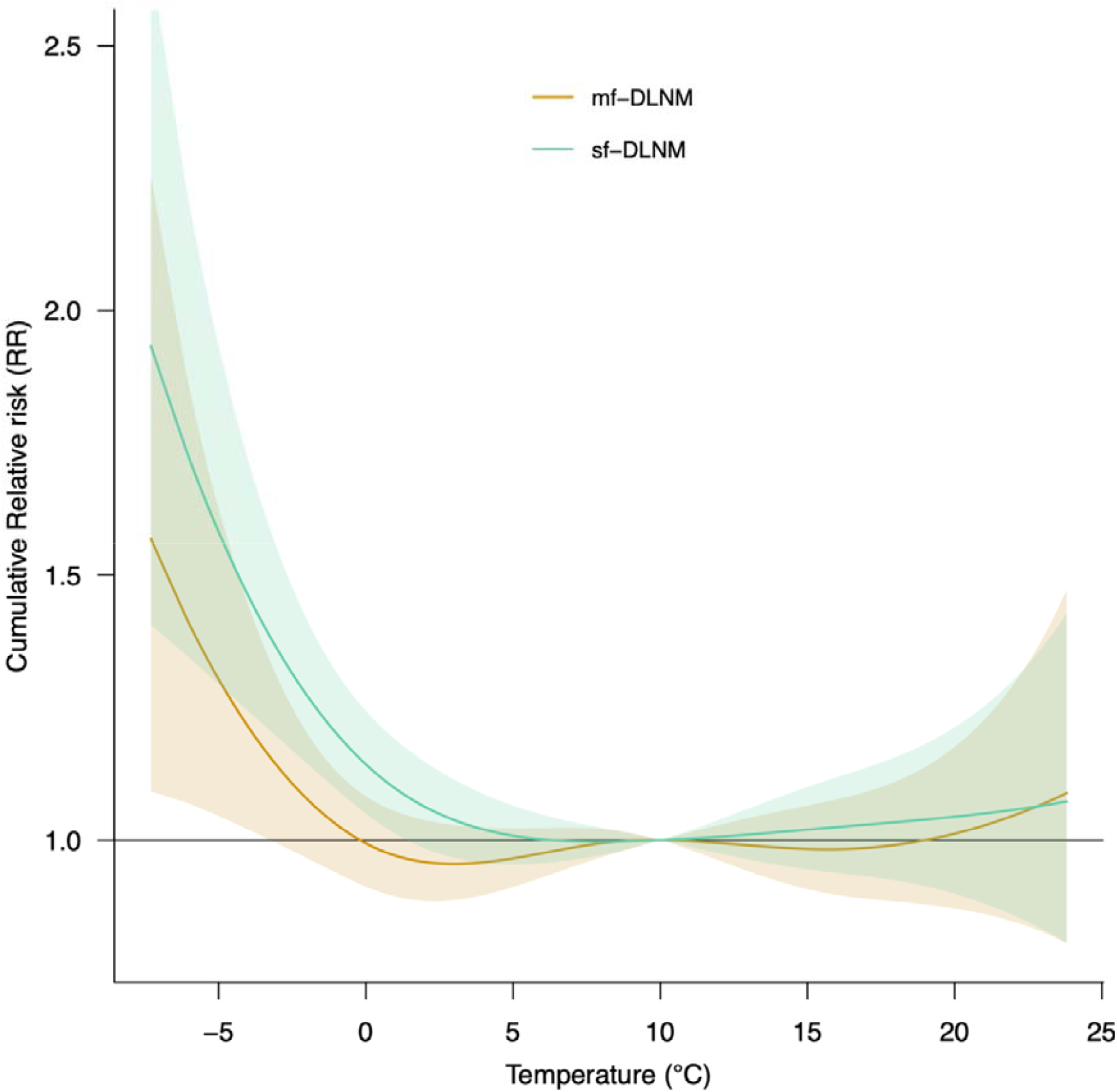
Overall cumulative relative risk (RR) of temperature on respiratory mortality over a 21-day lag, with a reference temperature of 10 °C, for the West Midlands, UK during January 01, 2010 to December 31, 2020 using mixed frequency DLNM (mf-DLNM).

**Figure 2.**
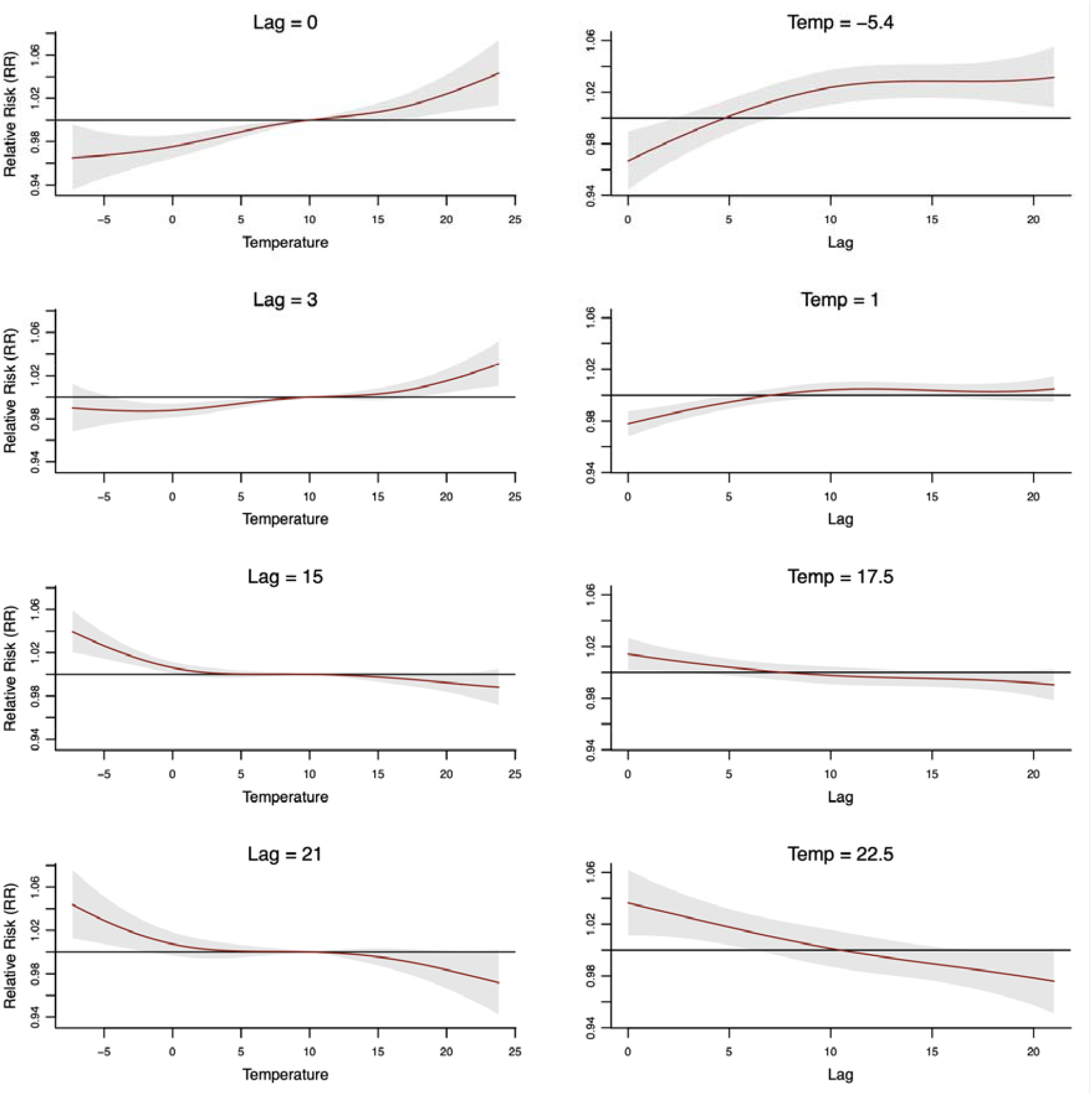
Relative risk (RR) of temperature on respiratory mortality in the West Midlands, UK during January 01, 2010 to December 31, 2020, at specific lags across temperatures (left), and at a specific temperature across lags (days) (right) using mixed frequency DLNM (mf-DLNM). The reference temperature is set to 10°C.

Furthermore, the predictive deviation between mf-DLNM and sf-DLNM was computed to assess model performance. **Table 2** presents the prediction deviations of overall RR of daily respiratory mortality associated with temperature percentiles between the mf-DLNM model and the sf-DLNM model. The absolute difference in the overall RR of mortality at −5.4°C (0.1^st^ temperature percentile) between mf-DLNM and sf-DLNM was 0.29, with a deviation of 0.26 and 0.32 for confidence interval predictions. The absolute difference in the overall RR of mortality at 22.5°C (99.9^th^ temperature percentile) between mf-DLNM and sf-DLNM was 0.00, with a deviation of 0.01 and 0.00 for confidence interval predictions. Therefore, if we take the estimated relationship from the sf-DLNM model as gold-standard, absolute performance deviation was observed to be higher for extreme cold temperatures, indicating a lower performance of the mf-DLNM in predicting cold-related effects. In contrast, the absolute performance deviation was smaller for extreme heat, suggesting that the model is more suitable for predicting the heat-related effect. Consequently, the mf-DLNM performed well in predicting the effect of extreme heat compared to extreme cold, indicating higher applicability for short-term studies, such as those predicting the health effects of heat waves.

**Table 2.** Overall cumulative relative risk (RR) at the 0.1st, 5th, 95th, and 99.9th percentiles of temperature for the mf-DLNM and sf-DLNM models, along with their respective absolute prediction deviations, for the West Midlands, UK during January 01, 2010 to December 31, 2020. Note. Reference temperature:10°C and Parentheses represent 95% CIs.

|  | mf-DLNM | sf-DLNM | Prediction Deviation |
| --- | --- | --- | --- |
| Temperature | RR | RR |  |
| -5.4 | 1.34 (1.06-1.71) | 1.63 (1.31-2.03) | 0.29 (0.26-0.32) |
| 1.0 | 0.97 (0.89-1.05) | 1.10 (1.01-1.19) | 0.13 (0.12-0.14) |
| 17.5 | 0.99 (0.89-1.10) | 1.03 (0.93-1.15) | 0.04 (0.04-0.05) |
| 22.5 | 1.06 (0.84-1.33) | 1.06 (0.85-1.33) | 0.00 (0.01-0.00) |

### 3.3 Method Performance Analysis

This section presents the results of the model performance evaluation based on performance metrics such as RMSE and the Pearson correlation coefficient (R). The RMSE values for the different models fitted to assess the mf-DLNM framework are shown in **Table 3**. The analysis indicated that mf-DLNM demonstrated performance comparable to that of the MIDAS model, underscoring its applicability to mixed-frequency datasets similar to those used in MIDAS. Furthermore, the performance difference between sf-DLNM and mf-DLNM was minimal, highlighting the potential of mf-DLNM approach in scenarios where high-frequency response variables are unavailable. **Figure 3** presents the scatter plots and Pearson correlations between weekly actual and predicted deaths from mf-DLNM and MIDAS. The figure shows that both models could predict the actual weekly death with nearly the same Pearson correlation coefficient (R), thereby affirming the utility of mf-DLNM for mixed frequency regression comparable to the MIDAS framework. However, estimating high-resolution health risks remains essential for environmental epidemiology, along with the relationship among mixed-frequency datasets. Therefore, a similar comparison was conducted between mf-DLNM and sf-DLNM for predicting daily deaths. **Figure 4** presents the scatter plots and Pearson correlations for predicting daily deaths from sf-DLNM and mf-DLNM. The R-value demonstrate that mf-DLNM can effectively estimate high resolution daily death, even in the absence of daily outcome data.

**Table 3.** The root-mean-square error (RMSE) for the predictive performance of mf-DLNM and MIDAS for weekly predictions, and of mf-DLNM and sf-DLNM for daily predictions of respiratory mortality counts in regions of England from January 01, 2010 to December 31, 2020.

| Region | Weekly prediction |  | Daily prediction |  |
| --- | --- | --- | --- | --- |
|  | mf-DLNM | MIDAS | mf-DLNM | sf-DLNM |
| West Midlands | 28.5 | 30.2 | 5.83 | 5.77 |
| East Midlands | 24.8 | 26.0 | 5.13 | 5.1 |
| East of England | 29.5 | 30.7 | 5.92 | 5.88 |
| London | 23.8 | 25.2 | 5.18 | 5.15 |
| South West | 29.0 | 30.6 | 5.82 | 5.78 |
| South East | 39.2 | 42.3 | 7.74 | 7.57 |
| North West | **39.0** | 41.1 | **7.51** | 7.41 |
| North East | **17.3** | 18.0 | **3.97** | 3.96 |
| Yorkshire and The Humber | **27.6** | 28.6 | **5.70** | 5.66 |

**Figure 3.**
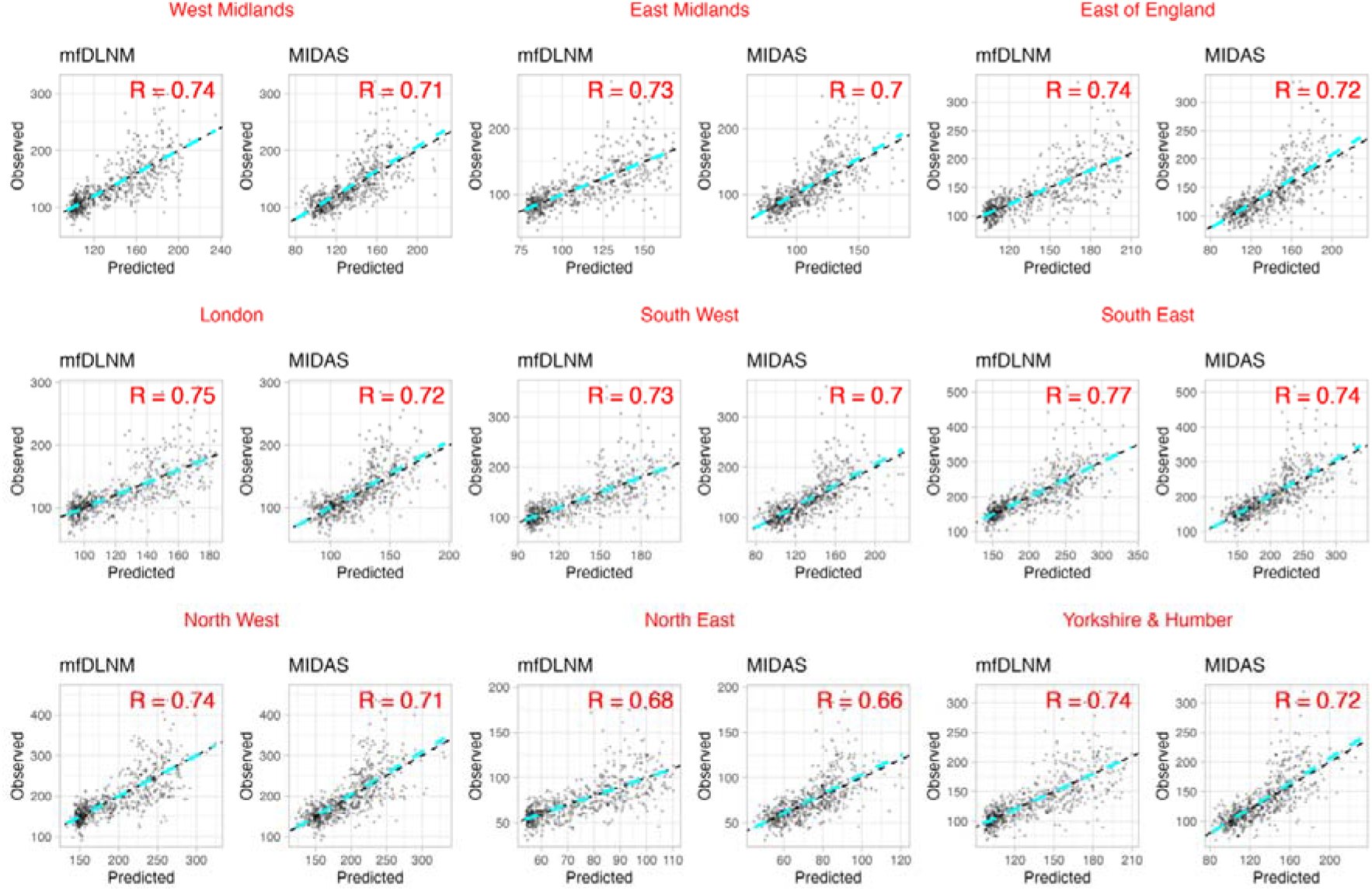
Scatter plots and correlation coefficients between weekly observed and predicted deaths by region in England. [left is mf-DLNM and right is MIDAS of the respective region]

**Figure 4.**
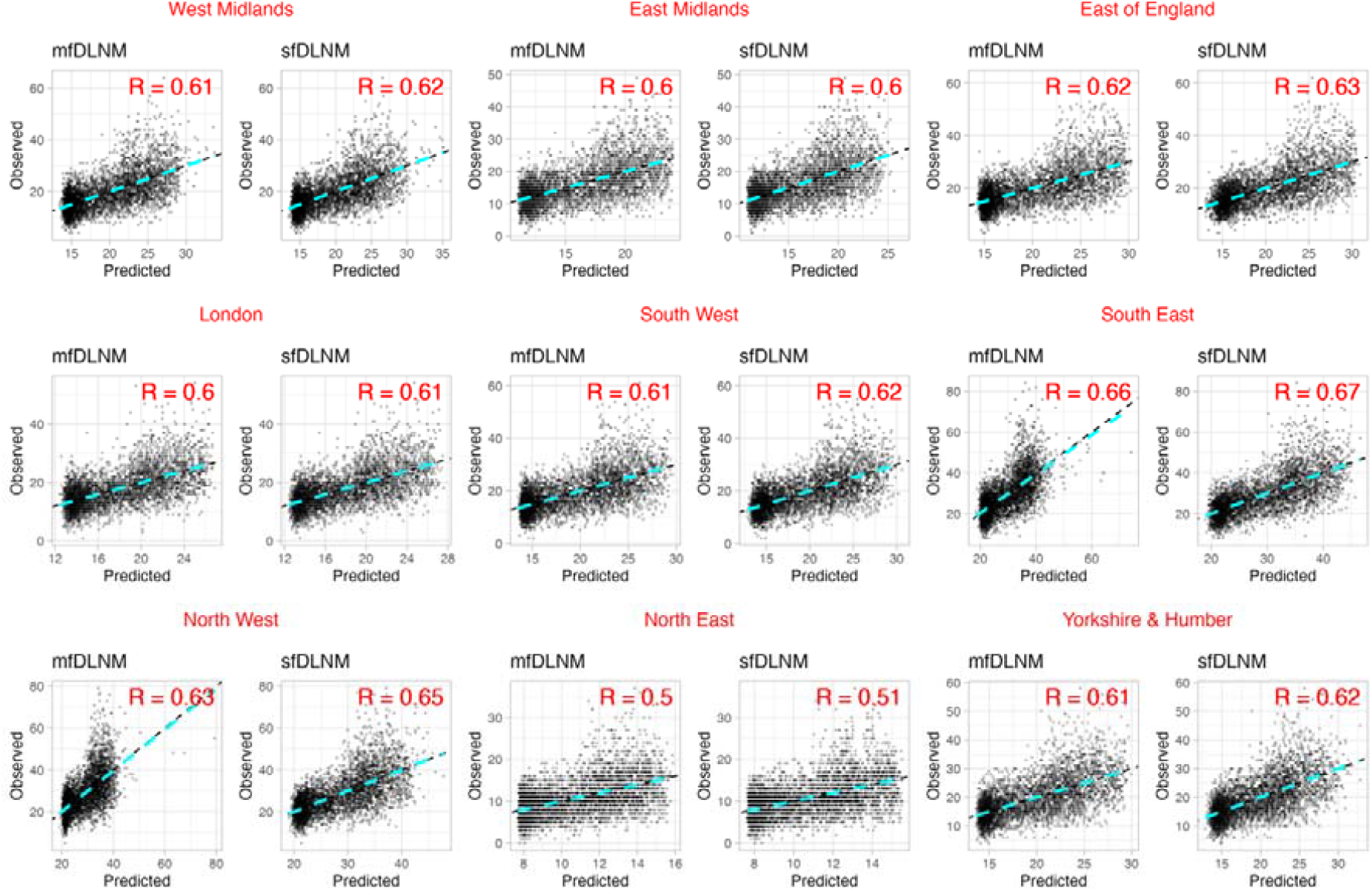
Scatter plots and correlation coefficients between daily observed and predicted deaths by region in England. [left is mf-DLNM and right is sf-DLNM of the respective region]

Across all regions, mf-DLNM consistently achieved predictive performance comparable to that of MIDAS and sf-DLNM. These findings highlight the robustness of mf-DLNM and its practical utility in real-world epidemiological applications. Therefore, the proposed mf-DLNM framework offers researchers the ability to utilize high-frequency exposure data without the need for temporal aggregation. As a result, even in the absence of daily health outcomes, high-resolution daily health risks can be accurately estimated using mf-DLNM.

### 3.4 Sensitivity Analysis

We conducted extensive sensitivity analyses to evaluate the impact of model choices. In particular, we evaluate changes in the model performance of the West Midlands region with varying degrees of freedom and lag dimensions in the models. We have also assessed the model sensitivity of different MIDAS specifications. **Table 4** presents the RMSE of weekly prediction (mf-DLNM and MIDAS) and daily prediction (mf-DLNM and sf-DLNM) using different model specifications, with an example of the West Midlands. The results indicate a slight acceptable change in the model prediction with changes in the degree of freedom and lag for all the models.

**Table 4.**
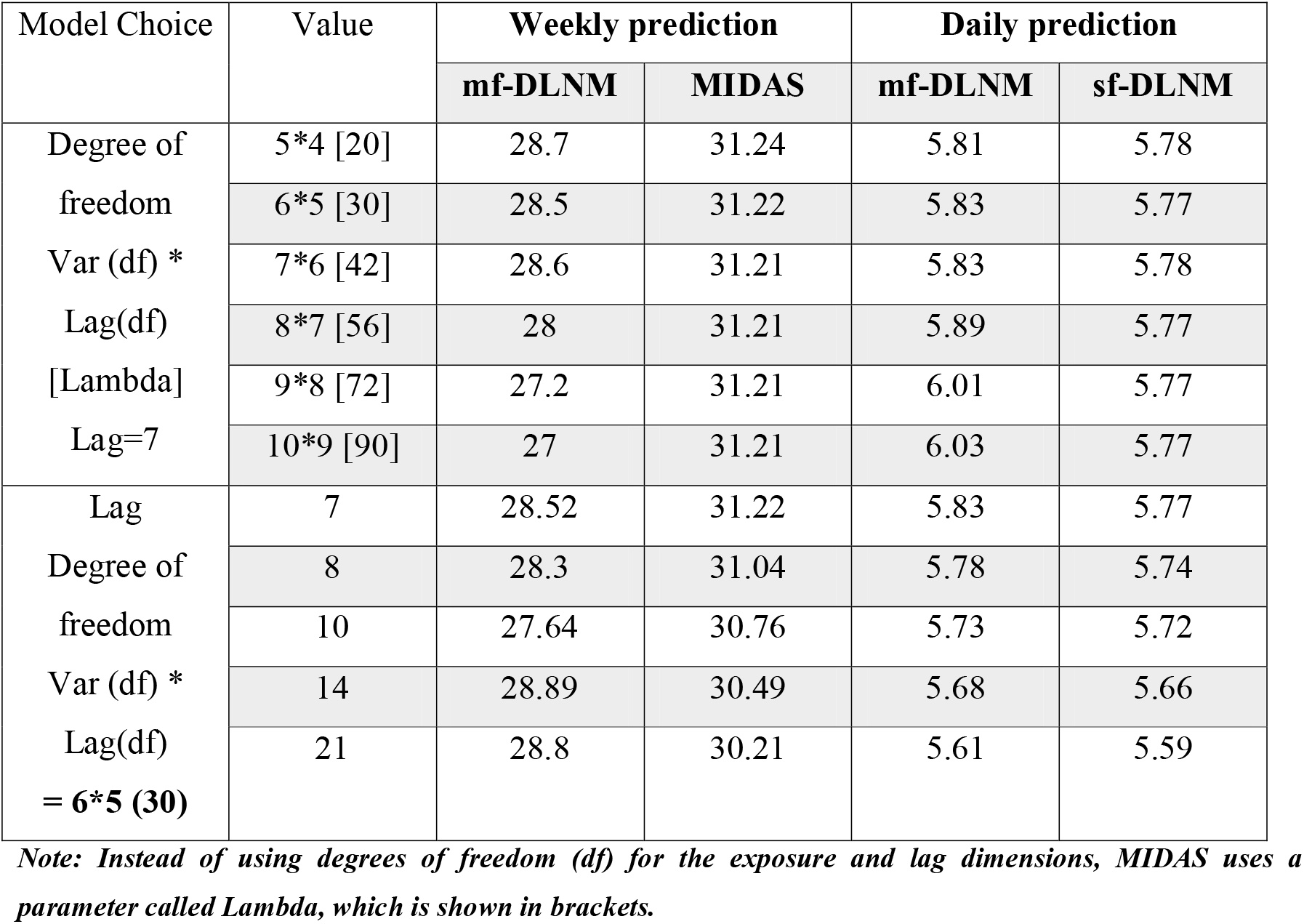
Sensitivity analyses of weekly predictions by mf-DLNM and MIDAS, and daily predictions by mf-DLNM and sf-DLNM, using root mean square error (RMSE) across different model specifications for the West Midlands example during 2010–2020.

The model sensitivity across different MIDAS specifications is illustrated in **Table 5**. The results indicate similar predictions across all MIDAS specifications. However, the non-Parametric MIDAS model performed best and was therefore used in the main analysis. For example in West Midlands, the RMSE for non-parametric MIDAS was 30.2, compared to 30.44 and 30.47 for nealmon and almonp MIDAS, respectively. This suggests lower sensitivity to these different MIDAS specifications. This suggests that the choice among these MIDAS specifications has minimal impact on predictive performance, implying low model sensitivity to specification.

**Table 5.** Sensitivity analysis of MIDAS model performance across regions of England (2010–2020) using root mean square error (RMSE) for weekly predictions of respiratory mortality counts under different model specifications.

| Region | Weekly Prediction |  |  |
| --- | --- | --- | --- |
|  | MIDAS<br>(nealmon) | MIDAS<br>(almonp) | MIDAS<br>(non-parametric) |
| West Midlands | 30.44 | 30.47 | 30.2 |
| East Midlands | 26.16 | 26.18 | 26.0 |
| East of England | 31.07 | 31.1 | 30.7 |
| London | 25.32 | 25.34 | 25.2 |
| South West | 30.69 | 30.72 | 30.6 |
| South East | 42.42 | 42.47 | 42.3 |
| North West | 41.41 | 41.43 | 41.1 |
| North East | 18.07 | 18.07 | 18.0 |
| Yorkshire and<br>The Humber | 28.88 | 28.92 | 28.6 |

## 4. Discussion

This study introduces the mixed-frequency regression approach for environmental epidemiology through the adoption of the mf-DLNM framework. Traditional methods, in the absence of high-temporal-resolution health outcome data, typically require the aggregating high-frequency exposure data to match the temporal scale of the outcomes. In contrast, the mf-DLNM eliminates the need to aggregate exposure data in environmental epidemiology studies, where exposure data are often available at higher time resolution than the outcome data. This approach enables researchers to use high-temporal-resolution exposure data and provides high-resolution risk predictions for informed public health decision-making.

The results of the demonstrative example, applying mf-DLNM to examine the relationship between temperature and respiratory mortality in the West Midlands, showed that the estimates from mf-DLNM were more consistent with those from sf-DLNM for extreme heat than for extreme cold. Therefore, this approach may be particularly useful for short-term environmental impact assessments, such as predicting excess mortality during heatwave events. Furthermore, the consistent model performance across nine regions with varying exposures and responses strengthens the evidence on the potential of mixed-frequency framework. This suggests that mf-DLNM can be a valuable alternative method for accurately estimating health risks associated with high-resolution environmental exposures, even in the absence of a high-resolution outcome data.

Moreover, there is a pressing need to advance these models, particularly in light of the increasing frequency of extreme weather events driven by climate change.^45^ In rare scenarios where high-resolution health data are available, studies have utilized these datasets to provide critical evidence on the impacts of exposures with high temporal resolution. For instance, a systematic review of thirty-three papers demonstrated that short-term air pollution exposures can have immediate and profound health effects.^46^ The review revealed that respiratory disease risk increases within 24 hours of exposure to PM_2.5_, PM_10_, and SO_2_. Similarly, other health conditions, such as an increase in cardiovascular disease risk, were observed within a few hours of exposure to high temperatures and various pollutants. These results underscore the importance of understanding the intricate and immediate connections between environmental exposures and human health. A study has shown that aggregating daily data to weekly levels introduces significant inaccuracies, particularly by underestimating the effects of extreme temperatures.^47^ Therefore, capturing short-term changes (hours to days) is crucial for exposure-response modelling, especially when accounting for highly variable environmental risk factors.^48^ While indicating the availability of high-frequency exposure information, researchers employ low-frequency exposure data for exposure-response modelling due to the unavailability of high-frequency response variables.^49^

As mentioned earlier, mixed-frequency modeling could resolve this challenge. MIDAS has been identified as a well-studied mixed-frequency regression model in econometrics.^50^ Additionally, it has been observed that MIDAS shares some similarities with DLM, which is utilized in environmental epidemiology for the development of DLNM. DLNM is a well-developed model with various crucial functionalities for environmental epidemiology. It is specifically designed for analyzing lagged and non-linear relationships, further, it has a robust and well-documented R package (dlnm).^17^ Furthermore, it integrates with Poisson regression, commonly used in epidemiology for count data such as disease incidences and its ability to incorporate GAMs provides more flexibility in modeling non-linear exposure-response relationships. However, MIDAS is not inherently designed to analyze non-linear predictor-response relationship. The visualization capabilities of DLNM, through the mentioned package, make it invaluable. Therefore, we developed mf-DLNM, inspired by MIDAS from econometrics, and employed the framework of DLNM to predict high-temporal-resolution risks using low-frequency response and high-frequency exposure data. Additionally, it is worth noting that this mf-DLNM can make high-frequency predictions of the response variable along with performing mixed-frequency regression, making the framework superior to MIDAS for environmental exposure modeling. Therefore, mf-DLNM would be suggested for the case where high-resolution risk estimation is required with a mixed-frequency dataset. The key strength of this study lies in the generalizability of the proposed model. Although the demonstrative analysis was conducted using specific predictors and responses, the mf-DLNM framework is flexible and can be extended to other exposures, health outcomes, and geographical regions. Similar to standard DLNM, the mf-DLNM can be employed in time-series regression analyses involving various predictors and responses within the field of environmental epidemiology.^26,51,52^ This approach provides researchers with the opportunity to use high-frequency exposure data to estimate health impacts effectively, even when the corresponding health data are available at lower temporal resolutions. This capability is crucial for addressing the growing public health concerns associated with environmental changes. It facilitates the use of high-frequency exposure information when available, providing more accurate and predictive insights for policy decisions.

We acknowledge certain limitations in our performance assessment methodology. The present work has not analyzed the model with multiple covariates. However, both MIDAS and DLNM are well-tested with multiple covariates, suggesting that mf-DLNM could perform similarly with multiple covariates.^17,53^ Additionally, while this study used daily-weekly mixed-frequency data, other temporal mixed frequencies, such as daily-hourly data, could be explored. Hourly concentrations of pollutants such as PM_2.5_, NO_2_, and O_3_, as well as weather parameters, are commonly available worldwide from ground-based and other sensor-based monitoring technologies.^54–56^ These higher-frequency datasets might behave differently and could challenge the model’s performance or computational efficiency. Furthermore, the risk estimates presented in demonstrative example of West Midlands are for methodological research purposes and are not intended to inform policy decisions. The risk estimates obtained may vary with the inclusion of other potential confounders.

In summary, this study introduces a novel methodological approach to using DLNM for mixed-frequency data and demonstrates its potential for employment in situations where high-frequency exposures are available for low-frequency responses. Future work aims to conduct a simulation study focused on evaluating the model performance specifically for inference testing and assessing the model behaviour under different scenarios, including varying sample sizes, lag structures, and other relevant factors. Future research could test the model’s adaptability to other temporal mixed frequencies, such as daily-hourly frequencies, model performance with multiple covariates, as well as its application for other exposures and health conditions, to further evaluate its robustness and generalizability.

## 5. Conclusions

We investigated the predictive performance of mixed-frequency modelling with the DLNM framework and conclude that mf-DLNM could be an effective method to use when high-frequency exposure data are available for use with low-frequency response data, eliminating the need to aggregate the exposure data. In addition, mf-DLNM can estimate high-frequency health risks to aid in preventive decision-making during extreme environmental events, for example to inform adaptation measures. The model should, however, be further evaluated using simulation studies to provide more insight into its parameter estimation accuracy, as well as examined using additional exposure datasets such as air pollution data, incorporating different mixed frequencies including hourly and daily exposure and health data.

### CRediT authorship contribution statement

**Nidhi Shukla:** Writing – original draft, Writing – review & editing, Visualization, Validation, Software, Methodology, Investigation, Formal analysis. **Suzanne E. Bartington:** Writing – review & editing, Supervision, Conceptualization. **Anna Hansell:** Writing – review & editing, Supervision, Conceptualization. **Tim CD Lucas:** Writing – review & editing, Supervision, Resources, Project administration, Methodology, Validation, Investigation, Funding acquisition, Data curation, Conceptualization.

## Data Availability

All data produced in the present study are available upon reasonable request to the authors.

---

**Supplementary Table 1:** Sensitivity analysis by AIC for different degrees of freedom in mf-DLNM, MIDAS, and sf-DLNM for West Midlands during the period 2010–2020.

| Degree of freedom | AIC |  |  |  |
| --- | --- | --- | --- | --- |
|  | Weekly prediction |  | Daily prediction |  |
|  | mf-DLNM | MIDAS | mf-DLNM | sf-DLNM |
| 5*4 [20] | 6736 | 3.04 | - | 25407 |
| 6*5 [30] | 6711 | 3.04 | - | 25393 |
| 7*6 [42] | 6723 | 3.04 | - | 25402 |
| 8*7 [56] | 6629 | 3.05 | - | 25400 |
| 9*8 [72] | 6513 | 3.05 | - | 25402 |
| 10*9 [90] | 6487 | 3.05 | - | 25400 |

*Note: Instead of using degrees of freedom (df) for the exposure and lag dimensions, MIDAS uses a parameter called Lambda, which is shown in brackets.*

**Supplementary Figure 1:**
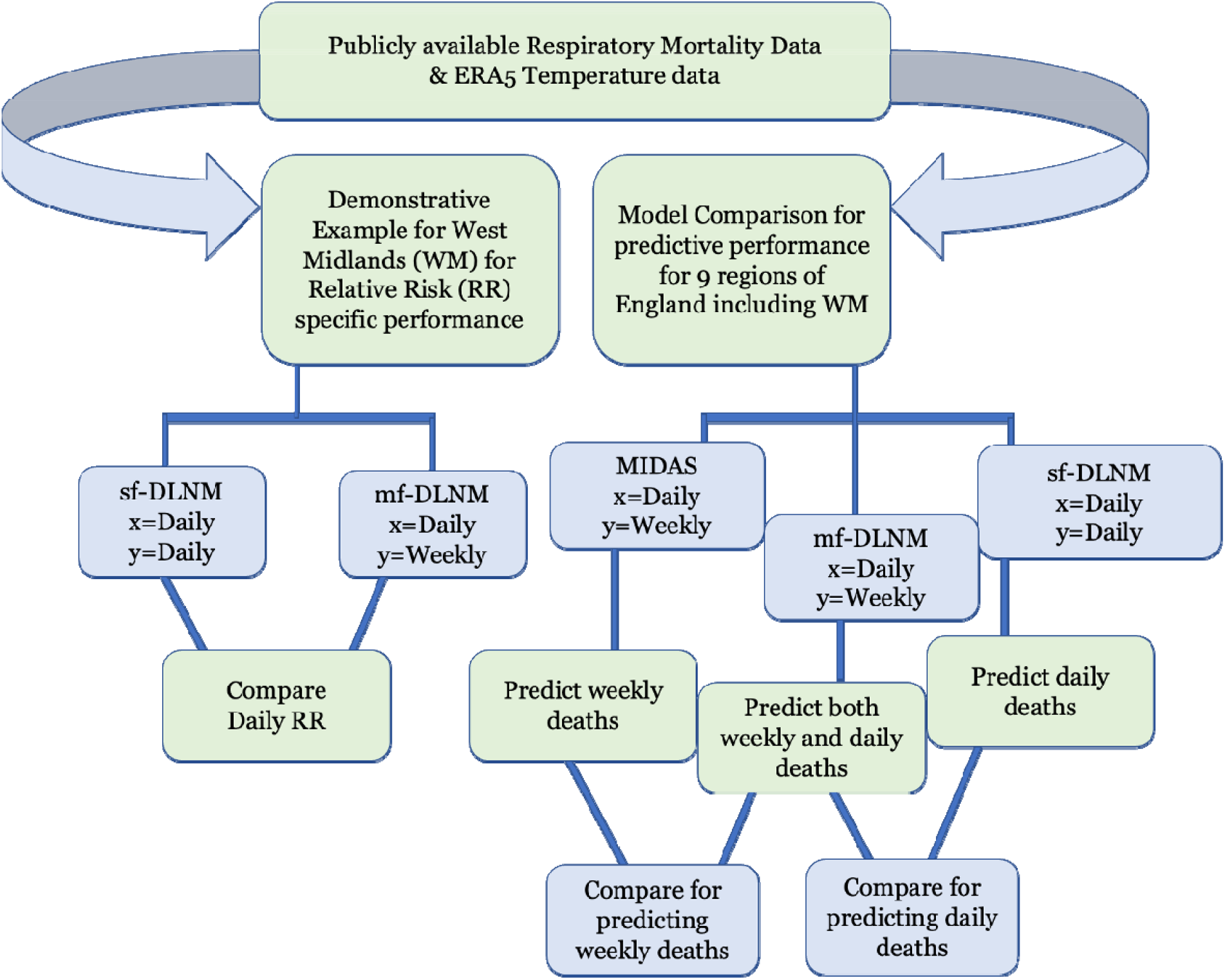
Workflow for evaluating mf-DLNM performance in predicting temperature-related respiratory mortality in England for mixed frequency dataset.

## Supplementary Example 1 (E1): Demonstrative example analysis

~~~
require(dlnm, dplyr, lubridate, mgcv, splines, ggplot2, Metrics)
# Load the chicagoNMMAPS dataset
data(chicagoNMMAPS)
#Weekly aggregation of outcome to test mf-DLNM
chicagoNMMAPS_w <-chicagoNMMAPS %>%
mutate(date = ceiling_date(date, unit = “week”, week_start = 7)) %>%
group_by(date) %>%
summarise(weekly_death = sum(death, na.rm = TRUE)) %>%
ungroup()
#merge by date daily and weekly for same exposure (temp)
chicagoNMMAPS_dw <-merge(chicagoNMMAPS, chicagoNMMAPS_w, by = “date”, all = TRUE)
# Model specification
var_fun <-“ns”; var_prc <-c(0.10,0.75,0.90)
var_knots <-quantile(chicagoNMMAPS_dw$temp, var_prc, na.rm = TRUE)
boundary_knots <-range(chicagoNMMAPS_dw$temp, na.rm=TRUE)
max_lag <-21; lag_knots <-logknots(max_lag, 3)
cen <-17
# Crossbasis
cb <-crossbasis(chicagoNMMAPS_dw$temp, lag=c(0,max_lag),
   argvar=list(fun=var_fun, knots=var_knots, Boundary.knots=boundary_knots),
   arglag=list(knots=lag_knots))
#fit the model mf-DLNM with weekly death
mfgam <-gam(weekly_death~ cb + ns(time,7*14), family=poisson(), chicagoNMMAPS_dw,
      method=‘REML’, na.action = na.exclude)
#fit the model sf-DLNM with daily death
sfgam <-gam(death~cb + ns(time,7*14),family=poisson(), chicagoNMMAPS_dw,
    method=‘REML’, na.action = na.exclude)
percentiles <-round(quantile(chicagoNMMAPS_dw$temp, c(0.001, 0.05, 0.95, 0.999), na.rm = TRUE),1)
#build crosspred for sf-DLNM and mf-DLNM
pred_sf <-crosspred(cb,sfgam, at=c(percentiles,chicagoNMMAPS_dw$temp), cumul=TRUE, cen=17)
pred_mf <-crosspred(cb,mfgam, at=c(percentiles,chicagoNMMAPS_dw$temp), cumul=TRUE, cen=17)
#####compare
comparison <-data.frame(
est_mf = pred_mf$allRRfit,
est_sf = pred_sf$allRRfit)
######
# Extract and combine results into tables
table <-rbind(with(pred_sf, cbind(allRRfit, allRRlow, allRRhigh))[as.character(percentiles),])
table1 <-rbind(with(pred_mf, cbind(allRRfit, allRRlow, allRRhigh))[as.character(percentiles),])
#ensure equal rownames/order
stopifnot(identical(rownames(table1), rownames(table)))
summary_table <-data.frame(
  Temperature = as.numeric(rownames(table1)),
  mf_RR = round(table1[, “allRRfit”], 2),
  mf_Low_RR = round(table1[, “allRRlow”], 2),
  mf_High_RR = round(table1[, “allRRhigh”], 2),
  sf_RR = round(table[, “allRRfit”], 2),
  sf_Low_RR = round(table[, “allRRlow”], 2),
  sf_High_RR = round(table[, “allRRhigh”], 2),
  Dev_RR = round(abs(table1[, “allRRfit”] - table[, “allRRfit”]), 2),
  Dev_Low_RR = round(abs(table1[, “allRRlow”] - table[, “allRRlow”]), 2),
  Dev_High_RR = round(abs(table1[, “allRRhigh”] - table[, “allRRhigh”]), 2))
##plot
pdf(“chicagoNMMAPS.pdf”, width=7, height=5)
col <-c(“darkgoldenrod3”, “aquamarine3”)
parold <-par(no.readonly=T)
par(mar=c(4,4,1,0.5), las=1, mgp=c(2.5,1,0))
plot(pred_mf, “overall”, ylim=c(0.8,2.5), ylab=“RR”, col=col[1], lwd=1.5,
  xlab=expression(paste(“Temperature (“*degree,”C)”)),
  ci.arg=list(col=alpha(col[1], 0.2)))
lines(pred_sf, “overall”, ci=“area”,col=col[2], lwd=1.5,
  ci.arg=list(col=alpha(col[2], 0.2)))
legend(“top”, c(“mf-DLNM”, “sf-DLNM”), lty=1, lwd=1.5, col=col, bty=“n”,
  inset=0.05, y.intersp=2, cex=0.8)
par(parold)
dev.off()
~~~

## References

1. Rocque RJ, Beaudoin C, Ndjaboue R, et al. Health effects of climate change: an overview of systematic reviews. BMJ Open 2021;11(6):e046333.

2. Arbuthnott K, Hajat S, Heaviside C, Vardoulakis S. Years of life lost and mortality due to heat and cold in the three largest English cities. Environ Int 2020;144:105966.

3. Sahani J, Kumar P, Debele S, Emmanuel R. Heat risk of mortality in two different regions of the United Kingdom. Sustainable Cities and Society 2022;80.

4. Vicedo-Cabrera AM, Scovronick N, Sera F, et al. The burden of heat-related mortality attributable to recent human-induced climate change. Nat Clim Chang 2021;11(6):492–500.

5. Zhao Q, Guo Y, Ye T, et al. Global, regional, and national burden of mortality associated with non-optimal ambient temperatures from 2000 to 2019: a three-stage modelling study. Lancet Planet Health 2021;5(7):e415–e425.

6. Momtazmanesh S, Moghaddam SS, Ghamari S-H, et al. Global burden of chronic respiratory diseases and risk factors, 1990–2019: an update from the Global Burden of Disease Study 2019. EClinicalMedicine 2023;59.

7. Wang YC, Sung FC, Chen YJ, Cheng CP, Lin YK. Effects of extreme temperatures, fine particles and ozone on hourly ambulance dispatches. Sci Total Environ 2021;765:142706.

8. Zhang Q, Peng L, Hu J, et al. Low temperature and temperature decline increase acute aortic dissection risk and burden: A nationwide case crossover analysis at hourly level among 40,270 patients. Lancet Reg Health West Pac 2022;28:100562.

9. Cui Y, Ai S, Liu Y, et al. Hourly associations between ambient temperature and emergency ambulance calls in one central Chinese city: Call for an immediate emergency plan. Sci Total Environ 2020;711:135046.

10. Ghysels E, Santa-Clara P, Valkanov R. The MIDAS touch: Mixed data sampling regression models. 2004.

11. Verbeek M. A Guide to Modern Econometrics John Wiley & Sons Ltd, 2004.

12. Gunay S, Can G, Ocak M. Forecast of China’s economic growth during the COVID-19 pandemic: a MIDAS regression analysis. Journal of Chinese Economic and Foreign Trade Studies 2020;14(1):3–17.

13. Karagoz K, Ergun S. Forecasting Monthly Inflation: A MIDAS Regression Application for Turkey. Turkish Journal of Forecasting 2020;4(1):1–9.

14. Yang C, Zhang R. Does mixed-frequency investor sentiment impact stock returns? Based on the empirical study of MIDAS regression model. Applied Economics 2014;46(9):966–972.

15. Basagaña X, Ballester J. Unbiased temperature-related mortality estimates using weekly and monthly health data: a new method for environmental epidemiology and climate impact studies. The Lancet Planetary Health 2024;8(10):e766–e777.

16. Gasparrini A, Armstrong B, Kenward MG. Distributed lag non-linear models. Stat Med 2010;29(21):2224–34.

17. Gasparrini A. Distributed lag linear and non-linear models in R: the package dlnm. Journal of statistical software 2011;43(8):1.

18. Koyck LM. Distributed lags and investment analysis. Vol. 4 North-Holland Publishing Company Amsterdam, 1954.

19. Almon S. The distributed lag between capital appropriations and expenditures. Econometrica: Journal of the Econometric Society 1965:178–196.

20. Braga ALF, Zanobetti A, Schwartz J. The time course of weather-related deaths. Epidemiology 2001;12(6):662–667.

21. Schwartz J. The distributed lag between air pollution and daily deaths. Epidemiology 2000;11(3):320–326.

22. Huang WTK, Braithwaite I, Charlton-Perez A, Sarran C, Sun T. Non-linear response of temperature-related mortality risk to global warming in England and Wales. Environmental Research Letters 2022;17(3).

23. Kephart JL, Sanchez BN, Moore J, et al. City-level impact of extreme temperatures and mortality in Latin America. Nat Med 2022;28(8):1700–1705.

24. Achebak H, Devolder D, Ballester J. Heat-related mortality trends under recent climate warming in Spain: A 36-year observational study. PLoS Med 2018;15(7):e1002617.

25. Chen R, Yin P, Wang L, et al. Association between ambient temperature and mortality risk and burden: time series study in 272 main Chinese cities. BMJ 2018;363:k4306.

26. Gasparrini A, Guo Y, Hashizume M, et al. Mortality risk attributable to high and low ambient temperature: a multicountry observational study. The lancet 2015;386(9991):369–375.

27. Lian T, Fu Y, Sun M, et al. Effect of temperature on accidental human mortality: A time-series analysis in Shenzhen, Guangdong Province in China. Sci Rep 2020;10(1):8410.

28. Sinko A. Some applications of mixed data sampling regression models. 2008.

29. Jacobson L, Oliveira BFA, Schneider R, Gasparrini A, Hacon SS. Mortality Risk from Respiratory Diseases Due to Non-Optimal Temperature among Brazilian Elderlies. Int J Environ Res Public Health 2021;18(11).

30. Mistry MN, Schneider R, Masselot P, et al. Comparison of weather station and climate reanalysis data for modelling temperature-related mortality. Sci Rep 2022;12(1):5178.

31. Roye D, Iniguez C, Tobias A. Comparison of temperature-mortality associations using observed weather station and reanalysis data in 52 Spanish cities. Environ Res 2020;183:109237.

32. Gasparrini A, Armstrong B, Kovats S, Wilkinson P. The effect of high temperatures on cause-specific mortality in England and Wales. Occupational and environmental medicine 2012;69(1):56–61.

33. Hajat S, Kovats RS, Lachowycz K. Heat-related and cold-related deaths in England and Wales: who is at risk? Occupational and environmental medicine 2007;64(2):93–100.

34. Ghysels E, Sinko A, Valkanov R. MIDAS Regressions: Further Results and New Directions. Econometric Reviews 2007;26(1):53–90.

35. Breitung J, Roling C. Forecasting inflation rates using daily data: A nonparametric MIDAS approach. Journal of Forecasting 2015;34(7):588–603.

36. Armstrong B. Models for the relationship between ambient temperature and daily mortality. Epidemiology 2006;17(6):624–31.

37. Ghysels E, Kvedaras V, Zemlys V. Mixed frequency data sampling regression models: the R package midasr. Journal of statistical software 2016;72:1–35.

38. Gasparrini A, Scheipl F, Armstrong B, Kenward MG. A penalized framework for distributed lag non-linear models. Biometrics 2017;73(3):938–948.

39. Hastie T, Tibshirani R. Generalized additive models: some applications. Journal of the American Statistical Association 1987;82(398):371–386.

40. Wood SN. Generalized additive models: an introduction with R CRC press, 2017.

41. Wood S, Wood MS. Package ‘mgcv’. R package version 2015;1(29):729.

42. Wickham H, Chang W, Wickham MH. Package ‘ggplot2’. Create elegant data visualisations using the grammar of graphics. Version 2016;2(1):1–189.

43. Li X, Zhang Y, Tian Z, et al. Lag effect of ambient temperature on respiratory emergency department visits in Beijing: a time series and pooled analysis. BMC Public Health 2024;24(1):1363.

44. Shao M, Yu L, Xiao C, et al. Short-term effects of ambient temperature and pollutants on the mortality of respiratory diseases: A time-series analysis in Hefei, China. Ecotoxicology and Environmental Safety 2021;215:112160.

45. Masselot P, Mistry MN, Rao S, et al. Estimating future heat-related and cold-related mortality under climate change, demographic and adaptation scenarios in 854 European cities. Nature Medicine 2025:1–9.

46. Wu K, Ho HC, Su H, et al. A systematic review and meta-analysis of intraday effects of ambient air pollution and temperature on cardiorespiratory morbidities: first few hours of exposure matters to life. EBioMedicine 2022;86.

47. Ballester J, van Daalen KR, Chen Z-Y, et al. The effect of temporal data aggregation to assess the impact of changing temperatures in Europe: an epidemiological modelling study. The Lancet Regional Health–Europe 2024;36.

48. Orellano P, Reynoso J, Quaranta N, Bardach A, Ciapponi A. Short-term exposure to particulate matter (PM10 and PM2. 5), nitrogen dioxide (NO2), and ozone (O3) and all-cause and cause-specific mortality: Systematic review and meta-analysis. Environment international 2020;142:105876.

49. Fang J, Song J, Wu R, et al. Association between ambient temperature and childhood respiratory hospital visits in Beijing, China: a time-series study (2013–2017). Environmental Science and Pollution Research 2021;28:29445–29454.

50. Ghysels E, Kvedaras V, Zemlys-Balevičius V. Mixed data sampling (MIDAS) regression models, Handbook of Statistics, Vol. 42. Elsevier, 2020.

51. Luo C, Qian J, Liu Y, et al. Long-term air pollution levels modify the relationships between short-term exposure to meteorological factors, air pollution and the incidence of hand, foot and mouth disease in children: a DLNM-based multicity time series study in Sichuan Province, China. BMC Public Health 2022;22(1):1484.

52. Vicedo-Cabrera AM, Sera F, Liu C, et al. Short term association between ozone and mortality: global two stage time series study in 406 locations in 20 countries. bmj 2020;368.

53. Franses PH. Yet another look at MIDAS regression. 2016.

54. Lang P. New approaches to the statistical analysis of air quality network data: insights from application to national and regional UK networks. University of York, 2020.

55. Schulte K. ‘Real-time’air quality channels: A technology review of emerging environmental alert systems. Big data & society 2022;9(1):20539517221101346.

56. Cromar KR, Duncan BN, Bartonova A, et al. Air pollution monitoring for health research and patient care. An official American Thoracic Society workshop report. Annals of the American Thoracic Society 2019;16(10):1207–1214.

